# Estimation of mRNA COVID-19 Vaccination Effectiveness in Tokyo for Omicron Variants BA.2 and BA.5 -Effect of Social Behavior-

**DOI:** 10.1101/2022.09.15.22280010

**Authors:** Sachiko Kodera, Yuki Niimi, Essam A. Rashed, Naoki Yoshinaga, Masashi Toyoda, Akimasa Hirata

## Abstract

Variability of COVID-19 vaccination effectiveness (VE) should be assessed with a resolution of a few days assuming that VE is influenced by public behavior and social activity. Here the VE for the Omicron variants (BA.2 and BA.5) is numerically derived for Japan’s population for the second and third vaccination doses. We then evaluated the daily VE variation caused by our social behavior from the daily data reports for Tokyo. The vaccination effectiveness for Omicron variants (BA.1, BA.2, and BA.5) are derived from the data of Japan and Tokyo with a computational approach. In addition, the effect of different parameters regarding human behavior on VE is assessed using daily data in Tokyo. The individual VE for the Omicron BA.2 in Japan was 61% (95%CI: 57%–65%) for the vaccination second dose from our computation, whereas that for the third dose was 86% (95% CI: 84%–88%). The individual BA.5 VE for the second and third doses are 37% (95% CI: 33%–40%) and 63% (95% CI: 61%–65%). The reduction of daily VE from estimated value was close correlated to the number of tweets related to social gathering in Twitter. The number of tweets considered here would be one of new candidates for VE evaluation and surveillance affecting the viral transmission.

## 1. Introduction

COVID-19 emergence in December 2019 caused significant mortality globally [1]. The pandemic became temporarily controlled in some countries owing to the development of new vaccines and its high vaccination rates [2]. However, resurgence has been observed, for example, in the latter half of 2021, in European countries and in Japan in early 2022. This would be mainly attributable to the Delta and Omicron variants’ higher infectivity [3] and the waning vaccination immunity [4]. The reduced vaccination effectiveness (VE) has also been reported extensively for the Omicron variant [5]. No restriction has been adopted in some countries after wide spread of variants in 2022.

In general, two measures are used to characterize vaccination related to the waning immunity: efficacy and effectiveness [6]. Vaccination efficacy is derived under ideal conditions, which does not always translate to real-world effectiveness. Thus, vaccination efficacy overestimates the effectiveness in practice. There are several mechanisms and factors that influence efficacy. One major issue is time-dependent antibody reduction, which is shown to be correlated with vaccination efficacy [7]. The neutralizing antibody kinetics was reported at 6 months following full vaccination with BNT162b2 [8]. Time dependent Nabs reduction, antibodies against the nucleocapsid, and the SARS-CoV-2 spike protein were measured for healthcare workers [9]. Observatory and computational studies suggested antibody reduction with time, which is a predictive metric of immune protection from symptomatic infection [10,11].

Effectiveness or real-world performance is rather divergent in different countries [6,12-14]. Higher protection levels against COVID-19 infection for the Delta variant were reported in a cohort study [12]; individual VE was 93% (95% CI: 85%–97%) in the first month after vaccination, but declined to 53% (95% CI: 39%–65%) after four months. The meta-analysis mean value for a systematic review of the Delta variant was 60.5% and 75.6% for partial and full vaccination, respectively [15]. In [16], Individual VE against symptomatic infection for the Delta variant was 89% (95% CI: 86%–92%) 7–59 days after a second dose, but declined to 80% (95% CI: 74%–84%) after ≥240 days. It increased to 97% (95% CI: 96%–98%) ≥7 days after a third dose. Instead, VE against symptomatic Omicron infection was 36% (95%CI: 24%–45%) 7–59 days after a second dose and almost no protection after ≥180 days. The VE then increased to 61% (95% CI: 56%–65%) ≥7 days after a third dose. In [17], the individual VE for Omicron variant BA.1 and BA.2 were almost the same against symptomatic disease, whereas 25 weeks or more after two doses, individual VE was 14.8% against BA.1 and 27.8% against BA.2. Booster immunization increased protection after a week to 70.6% against BA.1 and 74.0% against BA.2, waning to 37.4% for BA.1 and 43.7% for BA.2 at 15 or more weeks after receiving the booster dose. The reasons for these differences are unclear. We derived that the waning immunity of vaccination is the individual VE for the Delta and Omicron variants in our previous study [18]: for the second dose, the Individual VE for the Delta and Omicron BA.1 variants (two weeks after the 2nd shot) were estimated as 93.8% (95% CI: 93.1%–94.6%) for whole Japanese population and 62.1% (95% CI: 48%–66%) for the population in Tokyo, respectively.

The gap between efficacy and effectiveness as well as the variability of different countries would be attributable to daily behavior (e.g., mask wearing and social gathering). Most observatory studies were evaluated in terms of elapsed time after full vaccination (second dose) or booster shot (third dose). The observation time resolution was a few weeks to months (e.g., 2 weeks, 2, 4, and 6 months after vaccination), in addition that the different vaccination timing for each individual. However, behavioral change may be observed in a time resolution of a few hours to several days; e.g., superspreading events (social gathering) for the former and New Year holidays, etc., which are often characterized by the 3Cs (Closed spaces, Crowded places, and Close contact). Thus, the real-world VE reported in observatory studies can be considered as time-averaged value for such combined factors. An additional factor is population VE from the transmission of viral variants, which is closely related to herd immunity [19,20] or antibody prevalence ratio [21]. It would be helpful to discuss prevention measures as well as individual VE variability in different countries if population VE can be estimated in a resolution of a few days.

In this study, the waning VE immunity has been evaluated for the whole population in Tokyo from December 23, 2021 to March 5, 2022, for the Omicron BA.1 variant and that in Japan from April 11, 2022 to July 31, 2022, for the Omicron BA.2 and BA.5. The waning effect and behavior change on VE for infection prevention has been numerically estimated for the population in Japan for the second and third doses. We particularly considered the number of tweets associated with the risk of infection as one of the metrics.

## 2. Materials and Methods

### 2.1. Study Design

We have conducted two computational experiments to derive the VE. The first experiment is the derivation of individual VE for BA.1, BA.2, and BA.5 variants derived from Japanese population using weekly data. The other is the investigation of daily behavioral factors that characterize the daily reduction of individual VE in Tokyo. As input data, three metrics were considered: i) the mobility at the transit stations, ii) the keywords of Twitter (social drinking, BBQ, and Karaoke), and iii) the nighttime population who stayed in the downtown area, including restaurants and bars.

Approximately 90% of individuals were vaccinated with the Pfizer BNT162b2; the remaining is the mRNA-1273 Moderna COVID-19 Vaccine among the vaccinated population for the first and second shots in Japan. Similarly, Pfizer BNT162b2 (58.8%) and mRNA-1273 Moderna (41.1%) are mainly employed, in addition to Novavax (0.1%) [22]. Due to low Novavax percentage, two mRNA vaccines are considered jointly assuming that their VE are almost the same. Tokyo started to report the classification of new daily positive cases into “2nd (or more) vaccinated” or “unvaccinated” [23]. Note that in the third doses, Modera and Pfizer were vaccinated regardless of the manufacturer of the first and second doses.

### 2.2. Computational Approach

In [18], we proposed the computational method to estimate the waning immunity of vaccination among the Japanese population. The statistical data needed for this approach were the daily number of vaccinated individuals and daily positive unvaccinated cases and fully vaccinated individuals (second dose). This has been extended to treat the third and fourth doses.

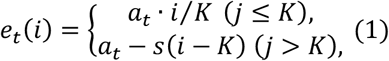

where *e*_*t*_(*i*) is the VE on *i*^*th*^ days after inoculation for *t*^*th*^ dose (= 1–4); and parameters of a and s were adjusted to reach a peak K days after inoculation (*K* = 14 for second dose, *K* = 7 for third and fourth dose), then decrease linearly. The individual VE for the first shot was assumed to be constant after 14 days due to a lack of data. As shown in Eq. (1), waning immunity is expressed as linear with time. This is consistent with the tendency of vaccination efficacy in [11] where neutralizing antibody is reduced with time almost linearly. Thus, for the same vaccine, this parameter s was kept as 0.15 (95% CI: 0.13–0.17) as in the second dose.

Population VE is required to estimate the effective unprotected population from infection. The Population VE *E*(*d*) was assumed to be as follows:

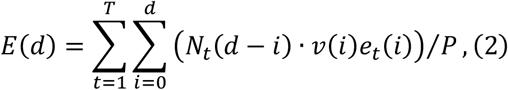

where *d* is the day index and *N*_*t*_ denotes the number of people who were newly administrated a vaccination dose (*t*= 1–4). P is the population of Japan (126,645,025 people) or Tokyo (13,843,329 people). The *v* denotes the SARS-CoV-2 sequences by variants as shown in Fig. 1a. The waning effect was overwritten when people took the second or third dose by adjusting the number of people vaccinated in the past [24].

**Figure 1.**
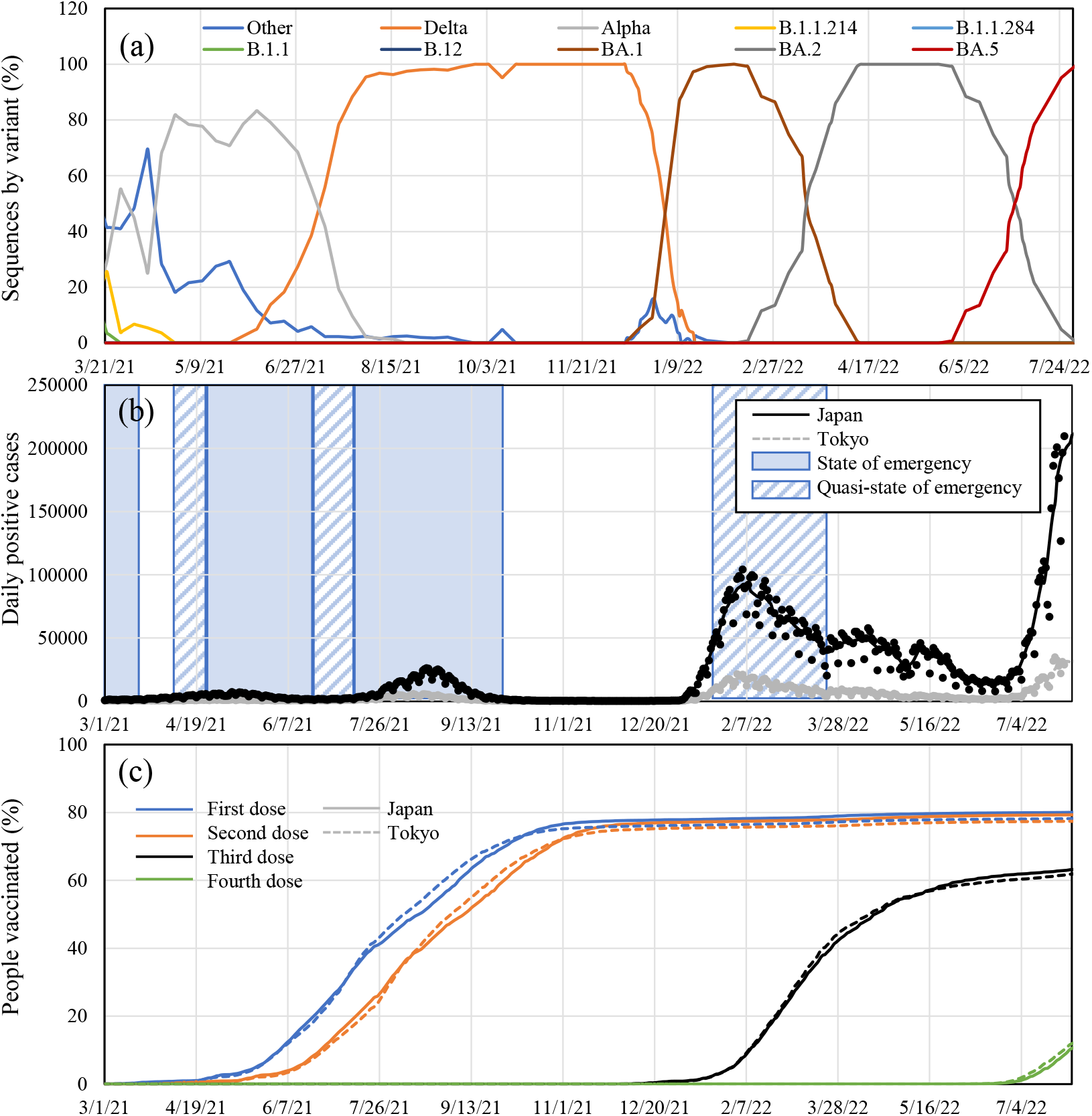
Time course of (a) SARS-CoV-2 sequences by variants in Tokyo. Time courses of (b) daily positive cases and (c) vaccination rate in Tokyo and over Japan.

An in-house python code was used to calculate the population VE. The optimal parameter at for each variant were derived by comparing the COVID-19 prevalence among unvaccinated people and without immunity to prevent infection. The weekly number of people without immunity to prevent infection was calculated from the 7-day average of daily population VE. The root mean squared percentage error (RMSPE) of that prevalence was used as an evaluation index (RMSPE < 10%).

### 2.3. Data for Vaccination

Vaccination in Japan began in March 2021, first for the medical workers, then the elderly individuals, and the June 2021 for non-medical workers, almost uniformly across the country. The interval between the first and second shots were controlled at 3 to 4 weeks. The third and fourth shot was then started from December 1, 2021 and May 25, 2022, respectively. The daily number of people who were newly administrated vaccination used in Eq. (2) are available from the Vaccination Record System (VRM) by the Digital Agency, Japan [25].

The data needed for fitting the parameter *a* for the *t*^*th*^ dose in Eq. (1) are from the website by the Ministry of Health, Labor, and Welfare [26] of Japan and the press release by the Tokyo Metropolitan Government [23]. The dataset in [26] includes the weekly number of unvaccinated, fully vaccinated individuals, and those vaccinated with booster dose, for the number of infected individuals in each category from April 11, 2022 to July 31, 2022. The Tokyo dataset for daily information [23], which is available only for Tokyo (population 13.9 million), will be used to discuss the effect of our behavior on VE. The overall age categories are not given for the Tokyo data. However, waning immunity is comparable to the age category >65 and <65 years in Japan (less than 2% after the second shot) [18]. Subjects without information regarding vaccination were excluded from this study (approximately 30%) [27]. The time slot from April 11, 2022 to May 25, 2022, and that from July 13, 2022 to August 7, 2022 were used for estimating VE for the Omicron variant BA.2 and BA.5, respectively. These correspond to the period >80% occupancy for each variant (See Fig. 1a). Effective vaccination of fourth shot reached less than 5%, considering a lag of one-week of effectiveness at the end of July 2022, the VE for fourth shot assumed to be marginal.

The SARS-CoV-2 sequences by variants in Tokyo are shown in Fig. 1. Note that the sequence for Japan is not available. Also, the number of daily positive cases and the percentage of people vaccinated are shown in the same figure.

### 2.4. Data Associated with Social Behavior

We considered three metrics considered to assess the effect of social behavior on the individual VE: i) mobility at the transit stations, ii) nighttime population who stayed in downtown area including restaurants and bars [28], and iii) Twitter key-words (social gathering for drinking and BBQ).

Mobility data [29] was often used as a metric for viral transmission, whereas it may not always be relevant to 3Cs as it may not clearly indicate the social behavior. We then considered the latter two metrics in addition to mobility.

The nighttime population who stayed in the area near restaurants and bars was obtained from NTT DOCOMO, INC, also accessible under the project of the Cabinet Secretariat COVID-19 AI Simulation Project, Japan.

Twitter data was used as it may indicate social activities where close contact oc-curs. The downtown population was considered as several domestic reports have in-dicated that the main infection clusters may be due to close contacts in these areas (see Fig. 2).

**Figure 2.**
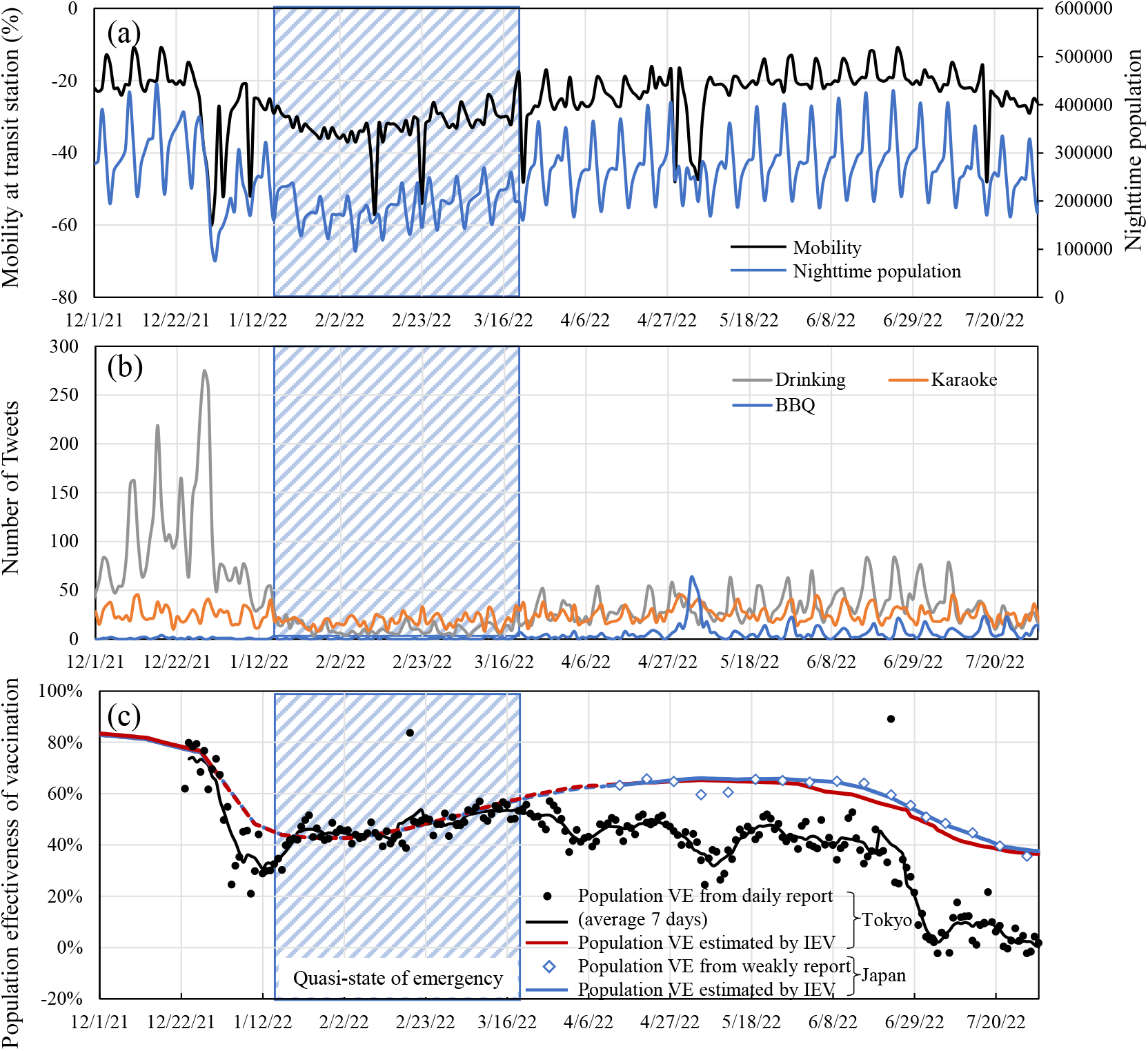
Time course of (a) mobility at transit stations and night population volume in downtown area of Tokyo, (b) the number of tweets for (i) social drinking and BBQ, and (c) Population VE estimated from daily report in Tokyo (dashed lines) and that weekly reported over Japan (solid lines).

Twitter data were obtained from NTT Data, INC. and processed by Toyoda Lab., University of Tokyo, and shared through the Cabinet Secretariat COVID-19 AI Simula-tion Project. Tweeted keywords completed during the day or the previous day, or those planned until the next day, were extracted when determining the number of tweeted keywords. Recorded data can clearly indicate time frames where these events are more popular while it is difficult to confirm if these gathering events are actually hold or not. For corresponding tweets, information on the prefecture was extracted from the user’s address.

A 7-day average value is considered considering the weekly effect when discuss-ing the correlation with VE. Also, an 8- and 5-day time lag on New Year holidays and golden week, respectively, which corresponds to the incubation and delay until the re-port for Delta and Omicron variants [30].

### 2.5. Statistical Analysis

To assess the effect of human behavior on the population VE, the statistical analysis was conducted using the Python 3.9.7. A linear regression model was used to fit the drop from estimated population VE as dependent variables for each independent variables: the number of tweets, nighttime population, and the mobility at the transit stations. The Spearman’s rank correlation test was also conducted. Statistical significance was accepted at *p* < 0.05.

## 3. Results

### 3.1. Vaccination Effectiveness for BA.2 and BA.5

In Table 1, the parameters of Omicron BA.2 and BA.5 variants parameters were derived for the Japanese population following the procedure in Sec 2.2. From Table 1, the BA.2 variant, individual VE just after the third shot (*a*_3_) was 86%, which is larger than that of the second shot of 61%. For BA.5, the estimated value was smaller than that of the BA.2 variant. The parameters of Omicron BA.1 variants were also derived from the data in Tokyo (*a*_2_ = 63 (95% CI: 61–64) and *a*_3_ = 85 (95% CI: 80–90), due to lack of data for Japan. The parameters of *a*_2_ and *a*_3_ value for BA.2 were comparable to those of BA.1.

**Table 1.**
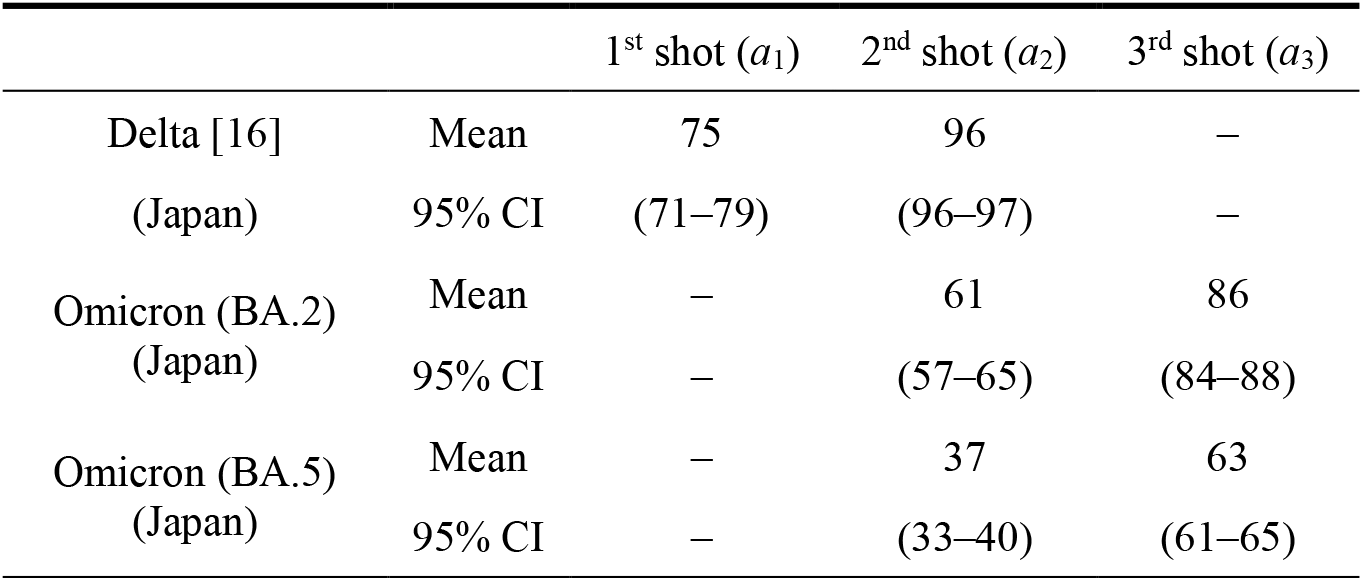
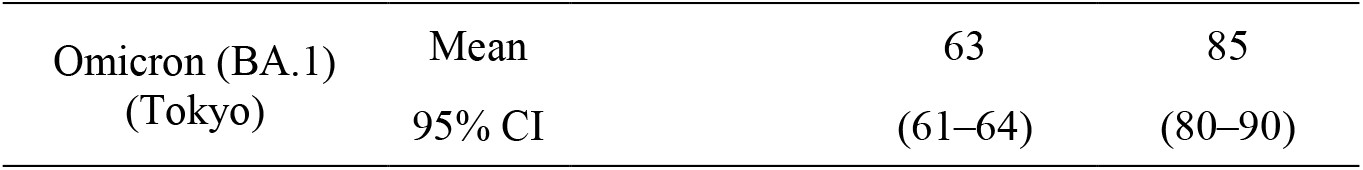
Parameters for the individual VE of second and third shots used in Eq. (1) for Omicron variants. For comparison, the parameters for parameters in Tokyo derived in our previous study [16] are also shown.

Population VEs over Japan (weekly) and Tokyo (daily) are calculated as shown in Fig. 2 (c) substituting the parameters derived in Table 1 into Eq. (2). The value calculated from the value reported by Tokyo is also plotted in the same figure. Similar tendencies are observed between the VE for the population over Japan and that in Tokyo. The former is 10%–15% higher than that of the latter averaged over 1 week from April to May (BA.2 variant) whereas 25%–30% in July (BA.5 variant). Clear decay is observed in some of the periods unlike in the 1-week averaged value.

### 3.2. Effect of Different Factors on Vaccination Effectiveness in Tokyo

Figure 3 shows the correlation between 7-day average daily drop from estimated population VE and different factors (mobility, Twitter, and nighttime population). The summation of drinking, karaoke, and BBQ were used as an index for Twitter. A strong correlation has been observed between them as shown in Figure 3. The slopes are different before and after the quasi-state of emergency. The correlation of the population VE reduction with Twitter including only drinking is after the quasi-state of emergency; R^2^ = 0.51 (*p* < .001), R^2^ = 0.12 (*p* = 0.01), and R^2^ = 0.71 (*p* < .001) before and after the quasi-state of emergency, and July, respectively. The Spearman’s rank correlation for each period is shown in Table 2. As the number of tweets, single keyword “social drinking” was considered (not shown in Fig. 3), in addition to the combination of three keywords (“social drinking”, “BBQ”, and “Karaoke”). Correlation was better for three keywords rather than single keyword of social drinking from March to May, 2022.

**Table 2.**
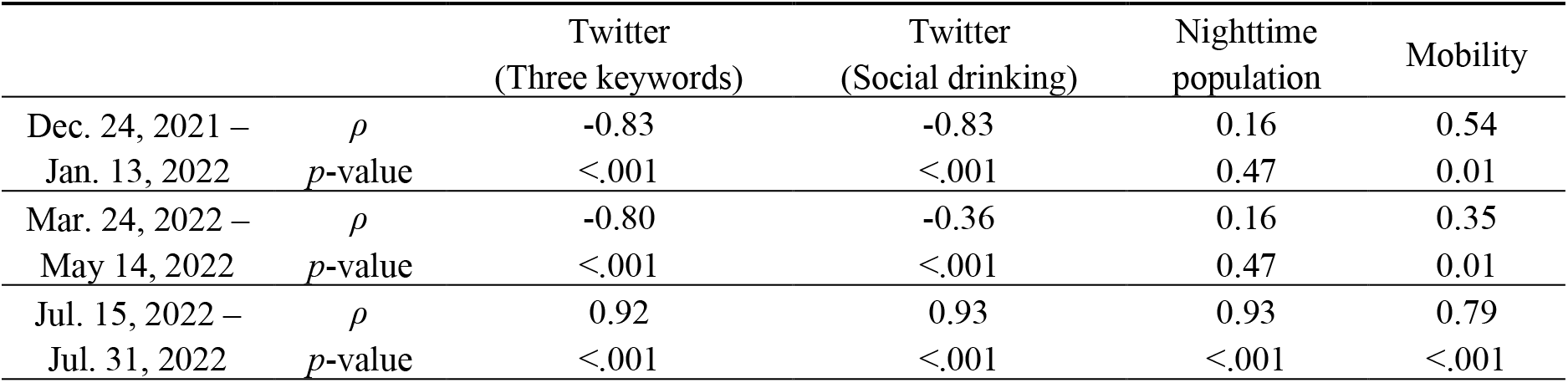
Spearman’s rank correlation between 7-day average daily drop from estimated population effectiveness of vaccination and different parameters of human behavior.

|  |  | Twitter<br>(Three keywords) | Twitter<br>(Social drinking) | Nighttime<br>population | Mobility |
| --- | --- | --- | --- | --- | --- |
| Dec. 24, 2021 – | $\rho$ | -0.83 | -0.83 | 0.16 | 0.54 |
| Jan. 13, 2022 | $p$-value | <.001 | <.001 | 0.47 | 0.01 |
| Mar. 24, 2022 – | $\rho$ | -0.80 | -0.36 | 0.16 | 0.35 |
| May 14, 2022 | $p$-value | <.001 | <.001 | 0.47 | 0.01 |
| Jul. 15, 2022 – | $\rho$ | 0.92 | 0.93 | 0.93 | 0.79 |
| Jul. 31, 2022 | $p$-value | <.001 | <.001 | <.001 | <.001 |

**Figure 3.**
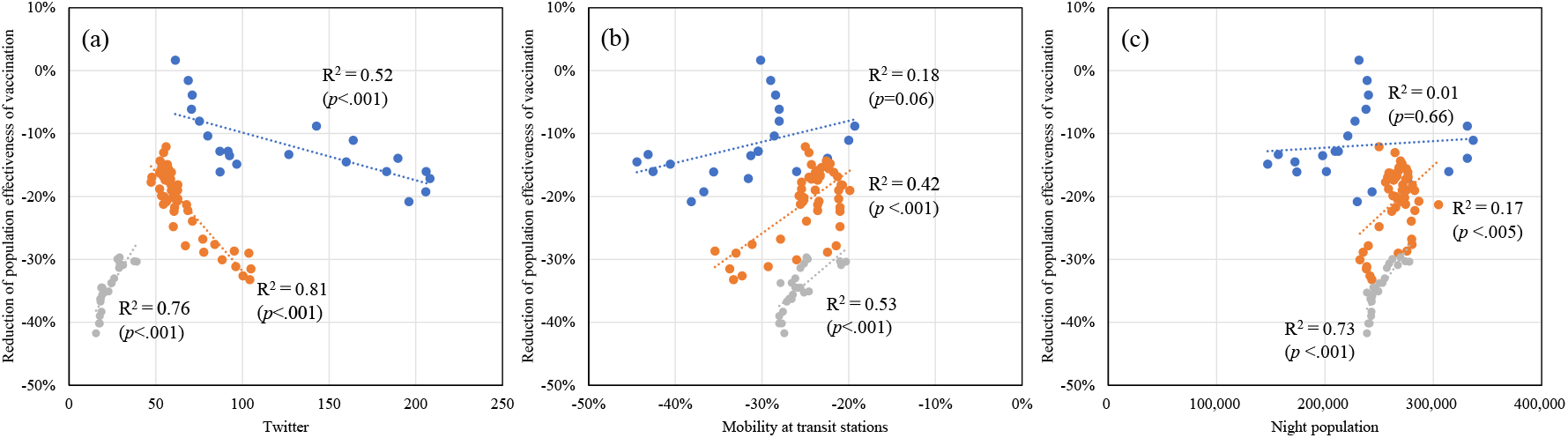
Correlation between 7-day average daily drop from estimated population effectiveness of vaccination and (a) Twitter, (b) mobility at transit stations, (c) nighttime population before and after the quasi-state of emergency. The period of blue plots, orange plots, and gray plots are from December 24, 2021 to January 13, 2022, from March 24, 2022 to May 14, 2022, and form July 15, 2022 to July 31, 2022, respectively.

## 4. Discussion

In this study, we numerically derived parameters characterizing the individual VE for Omicron variants: BA.1 for the population in Tokyo of 13.8 million and BA.2 and BA.5 for the population in Japan of 126.6 million. The feature of our approach ex-tended here based on our previous study [18] is that from a limited observatory data, the individual VE can be estimated with simple computation. In [18], the parameters characterizing VE, which are derived for the data from January 11, 2022 until January 24, 2022, worked well until early January.

We compared with the result in cohort study in Tokyo and suburb area [31] suggested individual VE for Delta and Omicron-dominant period was 88% (95% CI: 82–93) and 56% (95% CI: 37–70) for 14 days to 3 months after second dose to confirm our numerical derivation. Individual VE averaged over 14 days to 3 months were 88% and 53%, which are in excellent agreement with our computational model.

Individual VE for the BA.2 and BA.5 periods were derived for the Japanese population. The population VE were derived and compared with the observed data in Tokyo. The individual VE of second dose BA.1 in Tokyo and BA.2 in Japan were 63% (95%CI: 61–64) and 61% (95%CI: 57–65) after 14 days. From Eq. (2), estimated individual VE for BA.1 and BA.2 after 25 weeks were 37%, which are in fair agreement with 14.8% against BA.1 and 27.8% against BA.2 [17]. In addition, the third dose VE was 85% (95% CI: 80–90) and 86% (95%CI: 84–88) for BA.1 and BA.2, respectively. These values were 70.6% and 74.0%, respectively, in [17].

Slightly smaller population VE was observed than that of theoretical value except during the quasi-state of emergency. As shown in Fig. 3, the number of tweets, which are related to the infection risk, are correlated with the decrease in the vaccination effectiveness Unlike the Twitter data, correlation was observed but not strong for mobility and nighttime population. This hypothesized that the sentiment to the write user’s behavior in Twitter may reflect the decrease of VE rather than the physical number of phenomena, which may not directly lead to infection if appropriate countermeasure was taken. In Fig. 3, we have shown the combined data including social drinking, karaoke, and BBQ. The correlation of the reduction of population VE with Twitter only for social drinking decreased after the quasi-state of emergency (R^2^ = 0.51 [*p* < .001] and R^2^ = 0.12 [*p* = 0.01] before and after the quasi-state of emergency, respectively).

Some drop from the estimated value was observed during the New Year, golden week (in early May), in addition to heat wave with a time lag of approximately 5 days. In particular, a substantial decrease from the theoretical value was observed from the period of June 28 to July 15, 2022, which approximately corresponds to the national elections (from June 23 to July 10, 2022). The tendency in July was different from other periods. The bottom appeared around July 5, 2022, which potentially relates to the heat wave when the maximum daily ambient temperature was above 35°C. This out-of-season heat wave may cause less air ventilation.

The ratio of people who have immunity by infection was marginal because of the number of people infected with the Delta and Omicron variants until December 17 were less than 1.5% of the total Tokyo population and 0.7% of Japan, respectively. We considered that the people infected without symptoms would be 4 times higher than that of the reported cases [32]. The impact on the individual VE is marginal before the end of June. However, that effect may influence VE or immunity during the seventh wave where the BA.5 predominant. However, its impact may not be negligible in the seventh wave (from the end of June) because 14% of the unvaccinated population was infected; considering the non-symptomatic infection, it would be higher than 70%.

The population VE during the quasi-state of emergency, when such tweets are almost nonexistent, would be a good metric for vaccination efficacy. The population VE in Tokyo is smaller than that over Japan, which could be attributed to the population density [33,34]. The role of population density has been in different countries [35].

In summary, the VE for mRNA COVID-19 vaccines was numerically estimated from the limited information for the entire Japanese population. In addition, the number of tweets in Twitter was found to correlate with the decrease of the VE. The VE highly influenced viral transmission and thus, tweet monitoring would be an important factor for forecasting spread.

## Data Availability

All data produced in the present work are contained in the manuscript.

## Ethics and Permissions

No ethical permissions are needed in this study (this is a numerical study with open data).

## Acknowledgments

This research was funded by COVID-19 AI Simulation Project under Cabinet Secretariat, Japanese Government. The authors are team members of COVID-19 AI Simulation Project under Cabinet Secretariat, Japanese Government. Preliminary results in this study have been presented in the meeting of COVID-AI Simulation Project under Cabinet Secretariat, Japan (weekly reports). We would like to thank the participants meeting for providing useful comments and data needed in this study.

